# Identifying Homeless Population Needs in the Emergency Department Using Community-Based Participatory Research

**DOI:** 10.1101/2020.05.23.20104463

**Authors:** Andrew Franco, Jonathan Meldrum, Christine Ngaruiya

## Abstract

**Background:** Annually 1.5 million Americans face housing insecurity, and compared to their domiciled counterparts are three times more likely to utilize the Emergency Department (ED). Community Based Participatory Research (CBPR) methods have been employed in underserved populations, but use in the ED has been limited. We employed CBPR with a primary goal of improved linkage to care, reduced ED recidivism, and improved homeless health care.

**Methods:** A needs analysis was performed using semi-structured individual interviews with participants experiencing homelessness as well as with stakeholders. Results were analyzed using principles of grounded theory. At the end of the interviews, respondents were invited to join the “CBPR team”. At CBPR team meetings, results from interviews were expounded upon and discussions on intervention development were conducted.

**Results:** Twenty-five stakeholders were interviewed including people experiencing housing insecurity, ED staff, inpatient staff, and community shelters and services. Three themes emerged from the interviews. First, the homeless population lack access to basic needs, thus management of medical needs must be managed alongside social ones. Second, specific challenges to address homeless needs in the ED include episodic care, inability to recognize housing insecurity, timely involvement of ancillary staff, and provider attitudes towards homeless patients affecting quality of care. Lastly, improved discharge planning and communication with outside resources is essential to improving homeless health and decreasing ED overutilization. A limitation of results is bias towards social networks influencing included stakeholders.

**Conclusion:** CBPR is a promising approach to address gaps in homeless health care as it provides a comprehensive view incorporating various critical perspectives. Key ED-based interventions addressing recidivism include improved identification of housing insecurity, reinforced relationships between ED and community resources, and better discharge planning.

**Contribution to the Literature:**

- Homeless patients represent a significant portion of emergency department visits, but their health outcomes are worse and their unique health needs are often unmet. In this study, we utilized Community Based Participatory Research (CBPR) to integrate a complex health system with its community partners in order to determine and prioritize those needs.
- CBPR has seen limited use with emergency department patients, and provides insight on the needs of marginalized populations.
- This paper presents a set of recommendations for improving homeless health in the emergency department and decreasing recidivism.

## Background

Homelessness presents a major public health crisis in the United States, captured by overuse of Emergency Departments (EDs) as the de facto source of health care for many without housing. There are more than 1.5 million people who are homeless or in emergency shelters every year, and roughly 554,000 people were homeless on a single night in 2017.(1, 2) Definitions of homelessness include those who chronically lack fixed or permanent housing, are intermittently homeless, and are at-risk for homelessness.(3-6)

There are several health disparities among homeless individuals when compared to domiciled counterparts, most concerning of which is an increased mortality rate.(7-11) Homeless patients have been found to have nearly twice the mortality rate compared with non-homeless cohorts after adjustment for age, sex, and prior hospitalization.(8) This disparity is poorly understood but has been attributed to alcohol and substance abuse, mental disorders, homicide, suicide, and liver cirrhosis.(7)

People experiencing housing insecurity represent a disproportionate and growing share of ED visits. ED utilization by homeless patients is three times the US norm(12) and has increased 80% over the last 10 years.(13, 14) Homeless patients are more likely to be “frequent users” (≥4 visits/yr) or “super users” (≥20 visits/yr) of the ED, and thus utilize more resources.(15, 16) Homeless patients have twice the number of ED evaluations(17) and are four times more likely to re-present to the ED within three days of a prior evaluation compared to their housed counterparts.(13) Patients presenting to the ED are more likely to be homeless or at risk for homelessness. At one urban ED, 14% of total patients were living on the streets or shelters and 25% were concerned about becoming homeless within two months.(18) Only 4% of frequent ED users are discharged with a plan that specifically addresses their housing.(15)

Several risk factors for disproportionate ED use include lack of housing, substance abuse, decreased education, chronic illness, hunger, and exposure to violence.(6,12) Homeless patients are more likely to present to the ED with injuries.(19) Substance abuse and mental health disorders are independent risk factors for both homelessness and for being a high-frequency user of the ED.(16)

A high rate of ED use creates strain on the healthcare system and can lead to ineffective care for these patients. Homeless patients have an average of 2.32 days longer length of stay, and approximately $1,000 increased hospital costs per discharge.(20) Researchers have called for the development of ED-based interventions that specifically address homeless patients’ needs, reduce mortality, improve cost-effectiveness, and relieve overcrowding.(15, 21)

One approach to address populations at risk is Community Based Participatory Research (CBPR). CBPR is an approach to research which places the target population at the center of the process in addressing their priority issues in close collaboration with key community members.(22) While CBPR has been used extensively in the public health field, it has seen limited use in the ED.(23, 24) The aim of this project is to identify needs and improve care for the homeless population in the ED utilizing CBPR, with the ultimate goal of improving health outcomes and decreasing ED recidivism.

## Methods

### Study Setting

Our study was conducted in New Haven, Connecticut, an urban city with a population of 130,000 people in northeast United States.(25) Compared to cities of similar size in the US, New Haven has a high burden of poverty, crime, and homelessness.(25, 26) The 2017 Point-In-Time count found 543 people in shelters or on the street on a single winter night.(26) This number has decreased from prior years similar to many other major US cities.(27)

Yale New Haven Health (YNHH) is the largest hospital system in Connecticut. Yale New Haven York Street Campus is the largest hospital in the system and provides the majority of emergency care for homeless patients in the community it serves. It has more than 1,500 inpatient beds, and its ED sees more than 100,000 visits per year. As a teaching hospital, most patients are evaluated by residents or Advanced Practice Providers with attending physician oversight. Social workers are present all day and care coordinators are available during regular business hours to help address wider social needs of patients. Yale New Haven is one of three EDs in the US that participates in Project ASSERT (Improving Alcohol and Substance Abuse Services and Educating providers to Refer patients to Treatment), a screening and intervention program which helps patients with drug-related visits access treatment programs, shelters, and social agencies.(28, 29) There is also a dedicated Crisis Intervention Unit in the ED which provides treatment and referral for mental health emergencies.

There are multiple homeless services available in New Haven County. There are two main shelters for adults. People experiencing food or housing insecurities can call a 24-hour hotline by dialing “2-1-1” to be connected with community resources. A medical respite program provides recuperative care for patients being discharged from the hospital, lowering public healthcare cost and reducing unnecessary hospitalizations.(23) Federally qualified health centers and other outpatient primary care clinics provide care to patients at reduced cost and on a sliding scale regardless of income or insurance. The Veteran’s Affairs Errera Community Care Center serves veterans struggling with mental illness, substance abuse, and homelessness. The Community Health Care Van provides free primary care and harm reduction services to patients on the streets.

### Study Design

We utilized Community Based Participatory Research (CBPR) framework with the objective of identifying opportunities and interventions to improve the care of homeless patients in the Yale New Haven Emergency Department. CBPR ensures accountability of public health interventions by addressing inequities in health status associated with the marginalized and poor.(22, 30) Research methodologies, prioritization of needs, design of interventions, measures of effectiveness, and targets that define success are developed collaboratively.(22, 30)

According to the initial stages of CBPR guidelines, we identified key stakeholders working with people experiencing homelessness in the New Haven community **(Figure 1)**. Stakeholders were initially selected via a literature search and then supplemented through input from gatekeepers such as senior community leaders, hospital administration, and recommendations from established experts in the field. Recruitment was further augmented by snowball sampling utilizing gatekeepers and stakeholders.(31, 32) Key informant interviews were performed, and engagement of key informants was ceased upon saturation of themes(33, 34). This was achieved through iterative review of individual interviews as they occurred. A waiver was obtained through the Yale Institutional Review Board since the study objectives address quality improvement.

**Figure 1:**
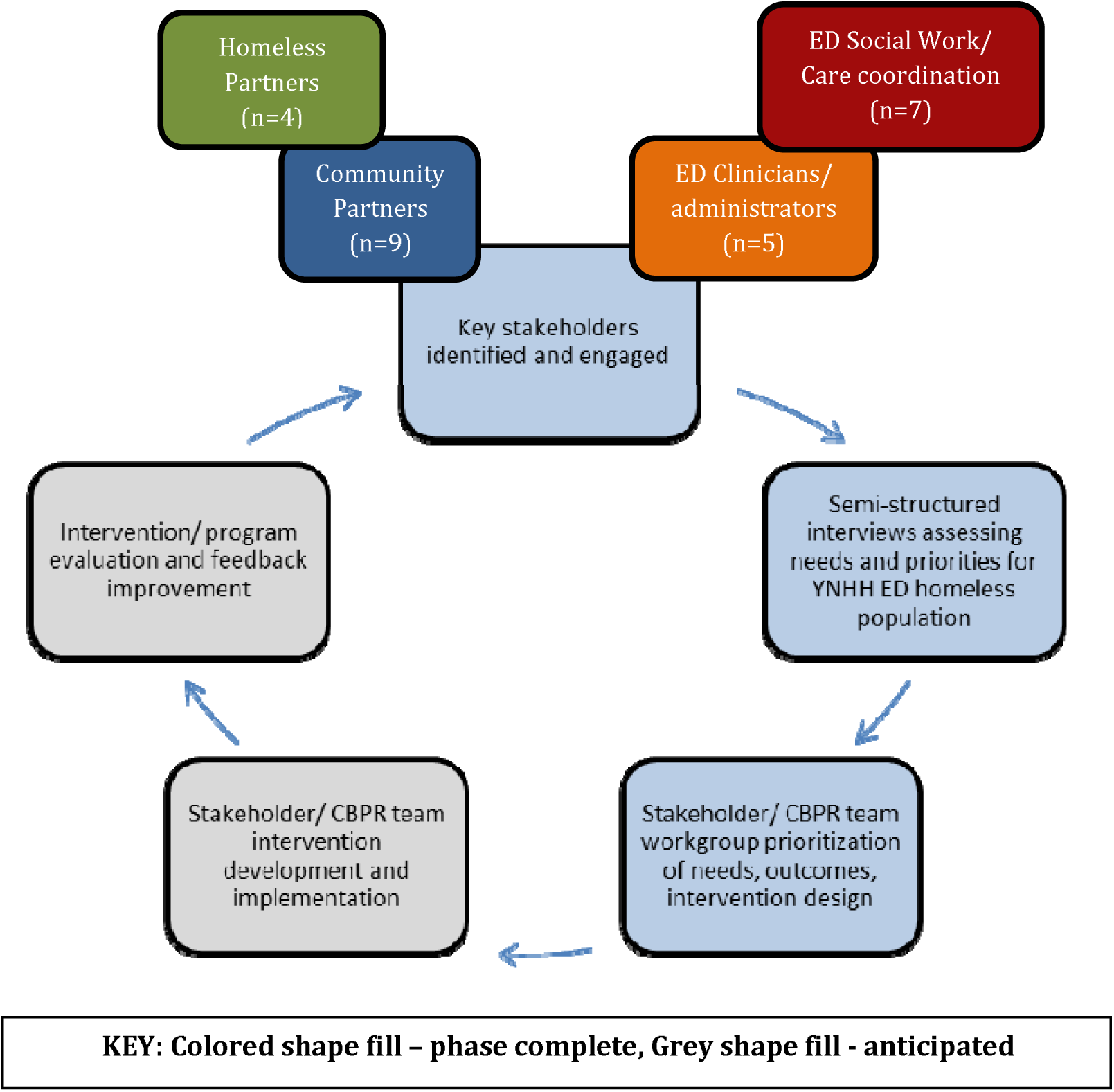
CBPR Schematic Chart.

One-on-one semi-structured interviews were conducted with each of the stakeholders privately by a single investigator from May 2016 to March 2017. Recording of interviews was offered in order to transcribe later, and in case it was declined notes were taken. Clients experiencing housing insecurity were interviewed at Columbus Respite Center. A standard interview guide **(Figure 2)**, was used to ensure that core concepts were addressed while still allowing for flexibility of conversation. Core concepts covered included descriptions of status quo, understanding barriers and limitations, identifying opportunities for improvement, and assessing willingness to participate in the CBPR team moving forward.

**Figure 2:**
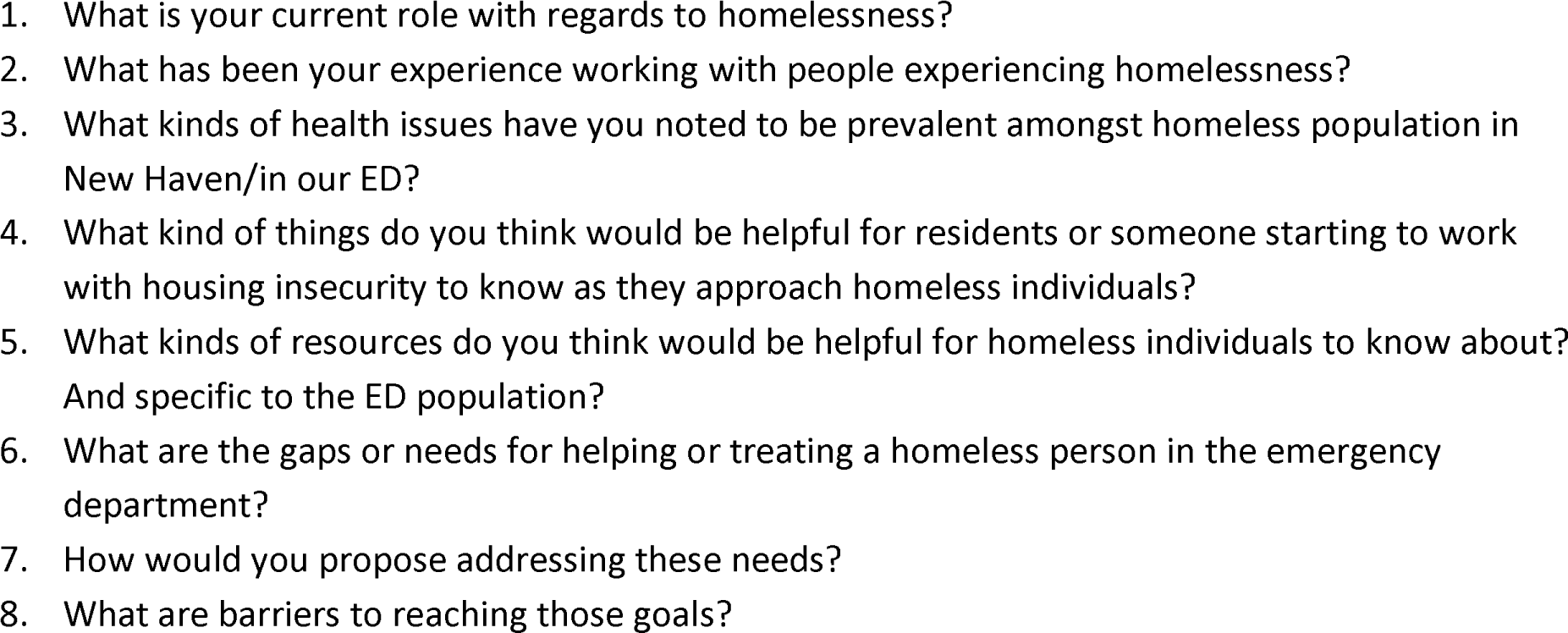
Semi-Structured Interview Guide.

The interview results were analyzed using grounded theory analysis,(33) themes were extracted, and these were used to build upon a flow diagram **(Figure 3)** to map interventions and affect outcomes for homeless patients treated in the ED.

**Figure 3:**
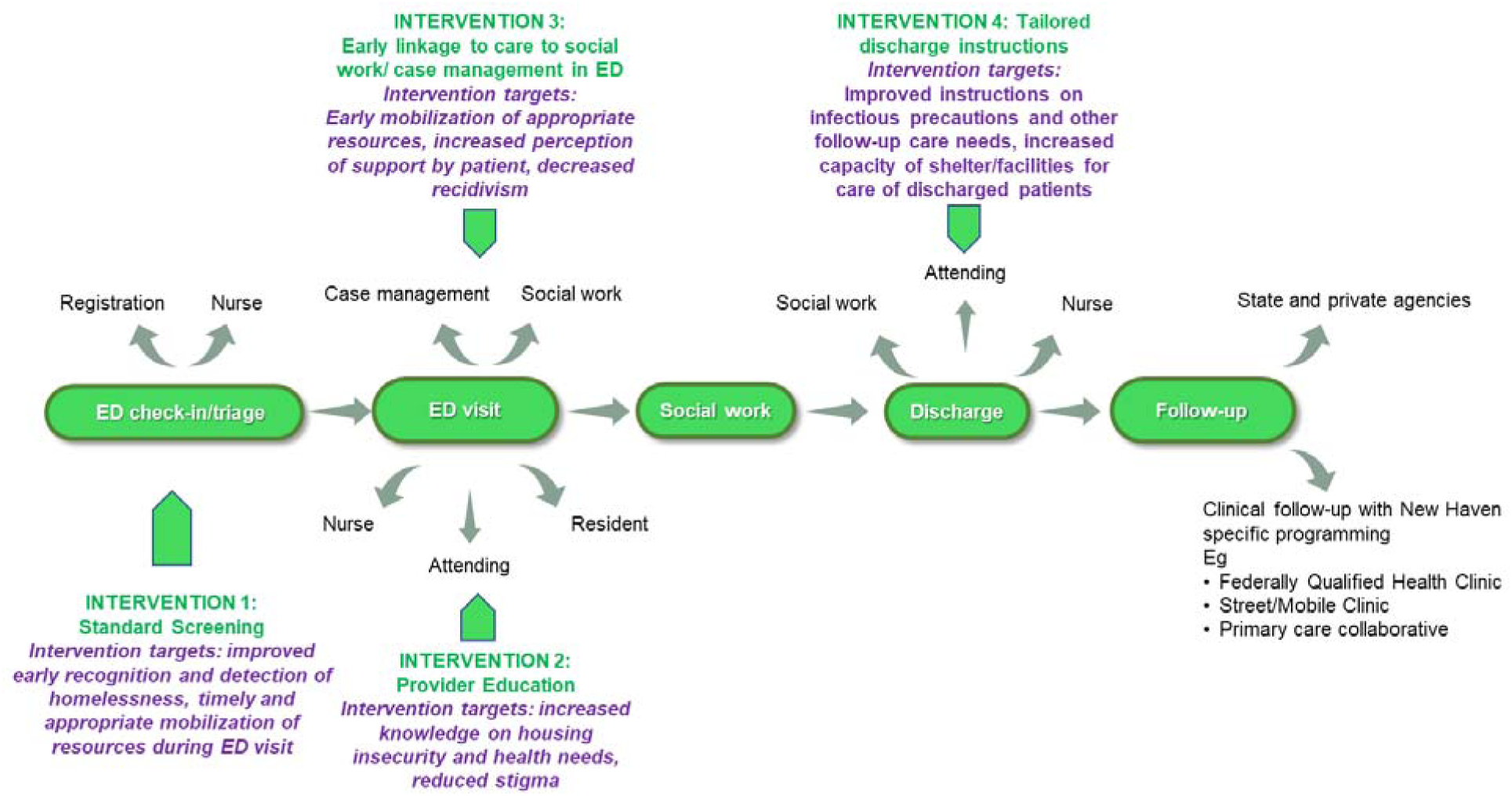
Flow Diagram of Planned Interventions.

At the end of each interview, all interviewees were invited to participate in the CBPR team **(Figure 4)**, and attend the CBPR team meetings which have continued on a monthly basis to date with intervention design and implementation (these results to be published elsewhere). The results from interviews were used to guide discussions during the CBPR team meetings, triangulating issues identified during the interviews, further identifying gaps and prioritizing interventions. In sum, the results of these interviews informed formation of the CBPR team, helped identify initial gaps for further exploration with the CBPR team once formed, and was key in developing relationships with community stakeholders.

**Figure 4:**
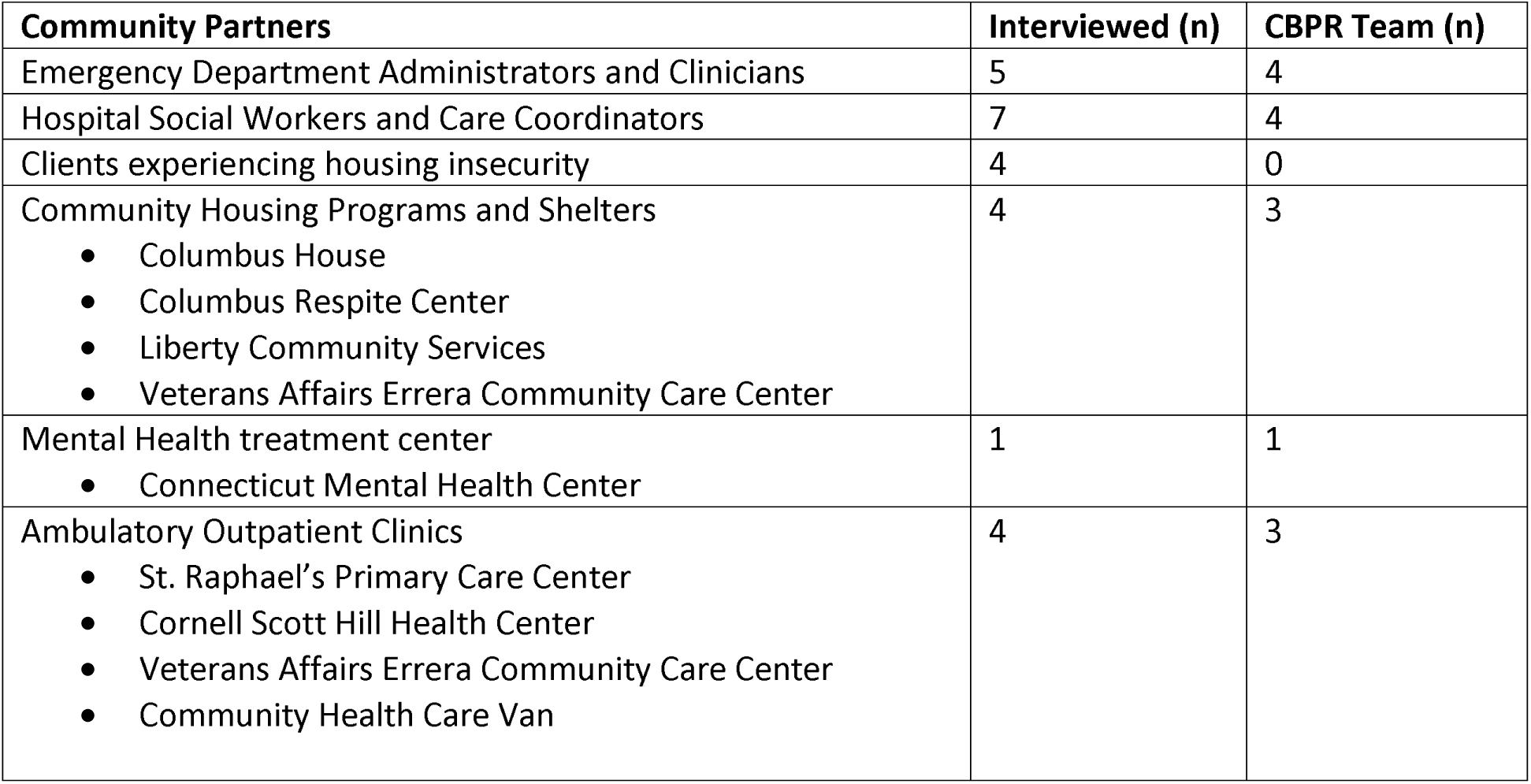
Community Partners.

## Results

Through CBPR methods we identified 25 community partners (**Figure 4**): 5 ED administrators and clinicians, 4 community housing programs and shelters, 1 mental health treatment center, 4 ambulatory outpatient clinics, 7 social workers and care coordinators in both the ED and inpatient services, and 4 clients experiencing housing insecurity. There was near equal breakdown by gender, multiple cultural backgrounds, and broad representation from various specialties and training, as well as levels of expertise in working with homeless. Analysis of the interviews revealed three recurring themes.

### Theme 1: The homeless population has a unique set of medical and social needs in the Emergency Department

#### There is a lack of access to basic needs, which often makes access to medical care needs insurmountable

Many interviewees discussed the difficulty of managing medical needs when basic social needs have not been met. One physician who provides primary care to homeless clients commented: “It’s about Maslow’s Hierarchy: it’s difficult to manage their complex medical conditions because they are focused on survival, food, shelter. You also have to focus on safety and security, even protecting their medications can be a challenge.”

#### Management of medical needs mandates concomitant attention to social ones for successful health outcomes

The social aspects of homelessness interact closely with medical needs. Homeless patients “can’t get better from even the most minor of illnesses: for example, injured extremities are very difficult to immobilize when you are constantly on the move”. Being homeless with an acute medical condition that usually could be managed at home may not be possible for a homeless patient: “There are a lot of patients we have no choice to admit because of medical problems that can’t be taken care of as an outpatient. For example, your patient has vomiting and diarrhea and doesn’t have reliable access to a bathroom.” Another spoke of the challenges of treating chronic medical conditions while homeless, “Basic self-care is challenging: we see lots of malnourished patients with poor control of their diabetes and hypertension.”

### Theme 2: There are specific challenges in addressing the needs of the homeless population in the Emergency Department

#### There is a lack of access to primary care, which results in episodic unsustained care

Despite a high proportion of the homeless population with insurance in New Haven, access to primary care is still a significant issue. An outpatient physician noted, “Even federally qualified health centers don’t have a unified focus and their services are too episodic, which is inadequate for this population.” As the door of the Emergency Department is always open this often acts as the main source of care for this population. One ED attending described the job of an ED physician: “Not just management of critically ill patients but also: risk stratification of undifferentiated symptoms, safety net for vulnerable populations (geriatric/homeless/substance/mental health), and coordination of care.” However other interviewees remarked that the systems within the Emergency Department are not designed to tackle a myriad of social needs.

#### Homeless patients are often unrecognized as such, which limits appropriate allocation of resources during their care

It is often not obvious that a patient is homeless or lacks secure housing. Multiple interviewees discussed that without identifying homelessness, clinicians will miss an essential component for their medical decision making and disposition planning. In the fast paced environment of the ED it is important that processes are in place to identify homeless patients. There was concern that patients “frequently provide former addresses or that of a trusted friend or relative in order to avoid stigma against homelessness”. This interacts with other systems including use of Electronic Medical Records, “Lynx had a section about housing. Epic doesn’t have this option, but can be placed as “homeless” under the problem list. However, this is often in flux, unlike hypertension or diabetes, so may not be up to date.”

#### Even when homeless patients are identified in the ED, the involvement of ancillary staff to assist with care needs is delayed

A common theme among social workers that we interviewed was the importance of early identification of homeless patients during their ED stay and integrating a social work assessment that begins alongside the medical workup. Delays in consulting social work can lead to suboptimal care. “Sometimes homeless patients are discharged to the waiting room and social work is called to address their needs while they are in the waiting room. What message is the Emergency Room sending to the patient?”

#### Provider attitudes and perceptions of homeless patients affect quality of care

One ED provider noted that stigma and anchoring bias may affect quality of care, and “we end up in a place of counter-transference, we get burnt out, and we start making fast cognitive decisions.” This can lead to providers being dismissive and missing dangerous diagnoses. Homeless clients described prejudices they felt from providers: “well Mr.-, why are you really here? You don’t really need to be here”, and that some staff members could be “snarky” and have “nasty attitudes”.

This can lead to distrust of care providers and missed opportunities for homeless patients to tackle the health issues they face during an ED stay. One staff member at a shelter described waiting “for the ‘ah-ha’ moment, where the patient will realize this is not what they want and make a change. This happens frequently.” This realization moment could be during an Emergency Department stay and by addressing mental health needs, substance abuse, and harm reduction providers can improve homeless health. “We don’t ask these patients enough: Do you want help with your substance abuse? Do you want to speak to a psychiatrist? There is a benign neglect that is not so benign of these very important issues. These patients often get short-changed in terms of their treatment and referral compared to their housed counterparts.” Another physician stated, “There are unique safety risks associated with being homeless; these patients lack an advocate, are not able to explain their illness clearly due to mental health and substance use, and we tend to view repeated ED visitation as protective, which I’m not certain is true. For example, homeless patient presents intoxicated with a head wound, later found to have brain bleed – likely will have delayed diagnosis of a serious illness.”

### Theme 3: Ensuring safe discharge planning and smooth care transitions to primary care and social services is essential

#### Improved discharge planning can curtail potentially avoidable ED visits

Discharge planning for homeless patients is more difficult than their housed counterparts. Investing the time to establish a plan for follow-up that is likely to be successful is challenging for a busy clinician, “The unaddressed needs of the homeless in the ED is the lack of follow up medical care.” One interview described the lack of a “long term plan” with patients being “discharged without clear discharge planning or adequate follow up”, which results “in a recurring cycle of returning to the ED.”

#### There is a need for improved communication on care plans with outside care services

One interviewee who works at a local shelter also commented on the challenges they face to support the health needs of recently discharged clients without health information. This interviewee shared a story of a patient diagnosed with a communicable disease who returned to a shelter without containment precautions, putting others at risk for exposure.

#### An improved system for linkage to care can improve utilization of available community resources

One ED physician felt that “getting homeless patients into primary care” is an essential goal in treating this population. The challenges of this were discussed by multiple people. One interviewee noted, “There’s no absence of medical resources, only a struggle to connect the resources with those who need them.” A homeless client agreed with this sentiment, “Certain resources were unknown to me until I was admitted to the hospital, when social work helped me out”. Connecting resources within the hospital setting with those in the community is an essential part of ensuring safe transitions of care. “The system doesn’t begin and end at the hospital door - there are a lot of services that need to be connected with.” As “there is no go-to person who treats homelessness in New Haven”, with multiple services and health care providers who work with the homeless, there lacks a unified pathway to guide homeless patients following hospitalization.

Multiple interviewees felt that traditional models of providing care through appointments does not apply, “their lives are so disorganized that we try to meet them on their own turf” and “handing a calendar of appointments doesn’t work for this group”. Integrating these concepts within the discharge planning process from the ED requires innovative thinking. One interviewee suggested open time slots in a primary care clinic that were also available for homeless patients after ED visits.

## Limitations

A possible limitation of our results is bias towards social networks influencing the stakeholders identified during interviews. Not all interviewees were able to commit to participating in the CBPR team with whom initial findings from interviews were further discussed. People experiencing homelessness were particularly unable to commit to regular meetings. This may have resulted in non-response bias. However, we felt this was addressed through a rigorous and iterative process incorporating a wide-range of sources to inform development of the CBPR team. Starting with a well-supported framework, we continually reassessed themes during the process of establishing the CBPR team.

## Discussion

We have described how Community Based Participatory Research can address the needs of homeless patients in the Emergency Department. To our knowledge our study is the first of its kind utilizing CBPR methodology to target homeless populations in the ED. By design, CBPR methods provide a novel approach to identifying gaps, designing interventions, and improving healthcare delivery to the homeless. The appropriate role of CBPR in identifying priorities for this population cannot be overstated, and the need to do so in a collaborative way with an interdisciplinary set of stakeholders is crucial to address complex needs.

Using semi-structured interviews, we identified three themes in the care for patients experiencing housing insecurity in the ED: (1) Homeless patients present with a unique set of medical and social needs, (2) There are specific challenges in addressing those needs, and (3) Safe discharge planning and transition to outside resources such as primary care and social services is essential.

Starting with the first theme, interviewees revealed that homeless patients require social needs to be addressed alongside medical ones. These social needs are complex but include food insufficiency,(35) dry clothing,(19) and shelter.(19, 23, 36) Consistent with prior research, our findings revealed significant overlap between homelessness and mental health disorders, alcohol use, and drug use.(12, 13, 37) These risk factors coexist with primary medical problems and prolong or complicate recovery and treatment, exacerbate homelessness and prevent effective social support or care management. Suggestions for improvement included enhanced connections between the ED, crisis intervention unit, and community substance abuse programs to build upon social support without investing excess effort duplicating services available outside the ED. This would require structured and regular communication between such outside resources. While this may require initial investments in administrative time, a more unified approach to addressing homeless needs is worthwhile.

Prior to our study, a partnership between Yale New Haven Hospital and Columbus House shelter formed to better serve homeless needs and decrease hospital utilization.(23, 24, 38) A prior CBPR study identified housing as a health concern not adequately addressed by health care providers upon discharge, and encouraged improved communication between hospital and shelter prior to discharge as well as a plan for safe transportation.(24, 38) Ultimately this partnership resulted in a medical respite for homeless patients, decreasing readmission rates and ED overutilization by providing a transitional facility for those with the greatest comorbidities.(23, 39) This includes nurse visits, meals, tobacco and alcohol cessation support groups, as well as housing placement services while in transition. This venture may be difficult to replicate in other settings. Our CBPR project built upon existing relationships between YNHH and Columbus House and included additional stakeholders of clinicians, case management, social workers, and other community actors. Furthermore, while an invaluable resource in our community, the respite almost exclusively admits patients from inpatient units not the ED and has a limited capacity. Our project focuses on additional contributors to the problem by focusing on early intervention and provider education, which has not yet occurred to date.

Our second theme revealed a set of unique challenges in treating homelessness, the first of which is identification. Knowledge of housing status affects diagnosis, disposition, and discharge planning, and thus was cited as a key place for improvement. Current methods of identification of housing insecurities in the ED were deemed inadequate by multiple respondents, and there is currently no screening tool being used. A prior study at our institution found that residents identify homeless patients by pattern recognition and stereotype such as poor hygiene or recurrent visits for alcohol use.(40, 41) This approach introduces bias and likely vastly underestimates the true burden of homelessness in the ED. Prevalence of housing insecurity in the ED varies by hospital and is poorly studied, but may be as high as 25% in some urban centers and is universally cited as higher than national estimates.(42, 43) Our findings suggest the need for universal screening in order to mobilize resources early in the ED visit to aid in disposition of the patient. One of our respondents also cited the success of regular screening in admitted hospital patients by the inpatient social work team which resulted in improved identification and resource mobilization. This or a similar screening process could be adapted for the ED.

Another challenge in addressing housing insecurity is provider bias. Several respondents who were health care providers cited a lack of clinical concern and burn-out as responsible for this trend. Similarly, our respondents experiencing housing insecurity noted feeling as though they were not treated as equals and felt that their providers were dismissive. All the same, there was a general consensus that prioritizing care for homeless patients is important. This stigma has been addressed in other studies, noting that there is no formal curriculum for educating providers about treating homeless patients(40) despite the American College of Emergency Physicians specifying that physicians-in-training should “recognize…socioeconomic status that may affect patient management” and lists “healthcare disparities” as a core competency in the practice of emergency medicine.(44) Curricula and clinical experiences have been used with mixed success to improve provider attitude towards homeless patients.(45-47) We propose instead that rather than lectures or increased homeless patient volumes, trainees may benefit from more humanizing interactions.(40) Examples of such training proposed by the CBPR team would include simulation scenarios or meeting with homeless patients outside of a clinical context in a service-learning environment.

The third and final theme relates to discharge planning and the unique challenges providers encounter when dispositioning homeless patients. It has been noted in prior work that coordinated transition from hospital to shelter may improve outcomes, and our research supports this finding.(23, 24, 38) CBPR discussion focused on decreasing ED recidivism through improved coordination of care. Prior literature suggests three potential intervention categories for reducing ED utilization: case management, individualized care planning, and information sharing.(48) Case management is an intensive individualized process whereby a single point of contact guides a patient through inpatient, outpatient, and community appointments.(49, 50) Individualized care plans are created at cross-departmental care plan meetings, however it has been noted that they are less comprehensive in their design and less effective than assigned case management.(48) The CBPR team discussed case management and individualized care plans as possible avenues to help navigate access to primary care and community assistance with housing, substance use, and mental health. An information sharing approach emphasizes shared case notes between providers in order to provide individualized care plans and improve ED overutilization.(51) This is most facile through the use of electronic medical records (EMR), however would be a challenge in New Haven as the YNHH EMR is separate from several of the federally qualified health centers. Instead, it was suggested that a HIPAA-compliant discharge sheet be printed and given to the patient to ensure safe transitions of care to outpatient providers and shelters.

Homeless patients represent a unique subset of the ED population at the interface of medical and social models of care. It has been noted before that the ED is the de-facto shelter and sobering center and serves not only as a medical but also social safety net.(41, 52, 53) There is a lack of adequate attention to homeless patients despite the significant resources allocated to this population in urban EDs across the US. The problem is a challenging one, given it requires multiple stakeholders, and comes against multiple competing interests in otherwise busy EDs.

A few factors may affect generalizability of our project. We recognize facilities and organizations supporting homeless needs in New Haven may be more abundant than other communities. Additionally, Yale New Haven Health is a large health system with many resources at its disposal. Ancillary support staff such as social work, care coordination, and Project ASSERT may not be available in every ED. 2-1-1 is a service that provides information for health, human, and social service across the United States and Canada, but assistance may not be as readily available in other countries.

We sought to outline specific opportunities with actionable interventions, as well as highlight representation from the community for which CBPR methodology was effective. We propose utilizing CBPR to enable interdisciplinary collaboration among key stakeholders managing homeless patients that interface with the ED. We are currently implementing interventions identified by the CBPR team, with whom indicators for success and best approaches for conducting our studies have been developed. The role of CBPR has been effective at identifying key needs in the homeless population in our ED, and we look forward to effective interventions to address recidivism.

## Data Availability

The authors confirm that the data supporting the findings of this study are available within the article [and/or] its supplementary materials.

**List of Abbreviations**
CBPR: Community Based Participatory Research
ED: Emergency Department
EMR: Electronic Medical Record
Project ASSERT: Improving Alcohol and Substance Abuse Services and Educating providers to Refer patients to Treatment
YNHH: Yale New Haven Hospital

## Declarations

A waiver was obtained through the Yale Institutional Review Board since the study objectives address quality improvement.

The datasets used and excerpts from performed interviews are available from the corresponding author on reasonable request.

There are no financial disclosures or conflicts of interest.

AF and CN conceived and designed the study. AF conducted the interviews and collected data. AF and JM analyzed the data and drafted the manuscript. CN provided oversight during drafting, wrote the study design section, and was involved with AF in the initial structuring and framing of the manuscript. All authors contributed to revision. AF takes responsibility for the paper as a whole.

## Acknowledgements

The authors would like to thank our community partners for sharing their time and experience. We also thank the ongoing efforts of the CBPR team in design and implementation of the described interventions.

